# Effect of intranasal insulin on osteocalcin levels and postoperative delirium in elderly patients undergoing joint replacement

**DOI:** 10.1101/2024.07.02.24309290

**Authors:** Yang Mi, Zhou Lei, Long Ge, Liu Xing, Ouyang Wen, Xie Chang, He Xi

## Abstract

**Background:** Recently, intranasal insulin has shown great promise in preventing perioperative neurocognitive disorders through ameliorating insulin sensitivity and cognitive function. Whether osteocalcin, as a bone-derived hormone that can directly regulate insulin sensitivity and cognitive function, is linked to the mechanism of intranasal insulin remains ill-defined.

**Aims:** To explore the effect of intranasal insulin on osteocalcin levels and the incidence and severity of postoperative delirium (POD) in elderly patients undergoing joint replacement.

**Methods:** The study is designed as a randomized, double-blind, placebo-controlled clinical study. 212 elderly patients (≥65) were randomly assigned to receive either twice 40 IU insulin (n=106) or placebo (n=106). The incidence and severity of POD were estimated by the Confusion Assessment Method (CAM) and the Delirium Rating Scale (DRS)-98. The levels of total osteocalcin (tOC), uncarboxylated osteocalcin (ucOC), insulin and glucose in venous blood and cerebrospinal fluid were detected by Enzyme-linked immunosorbent assay (ELISA). Insulin sensitivity was assessed by Homeostasis model Assessment of Insulin Resistance (HOMA-IR). The primary objective was to compare the difference of osteocalcin levels and insulin sensitivity between two groups, with the secondary objective to compare the difference of POD incidence and severity.

**Main Results:** It showed that 8 patients (8.33%) occurred POD in insulin group within 5 days after surgery, significantly fewer than 23 patients (23.23%) in placebo group (P=0.004). Mean peak DRS in insulin group was significantly lower than that in placebo group (P<0.001). After intranasal insulin intervention of 3 days, levels of tOC and ucOC in cerebrospinal fluid were significantly elevated in insulin group at D_0_ (all P<0.001). Levels of tOC in plasma were significantly higher in insulin group than that in placebo group on D_0_, D_1_ and D_3_ (all P<0.001). Plasma ucOC level in insulin group was higher on D_0_, but lower on D_1_ and D_3_ than placebo group (all P<0.001). HOMA-IR was significantly lower on D_3_ in insulin group than placebo group (P=0.002).

**Conclusions:** Intranasal insulin notably reduced the incidence and severity of POD in elderly patients undergoing joint replacement, and alco significantly improved central and peripheral osteocalcin levels and peripheral insulin sensitivity. Though these preliminary results needed further confirmation, it suggested that osteocalcin was promisingly involved in the mechanism of intranasal insulin in improving insulin sensitivity and POD.

**Trial registry numbers:** Chinese Clinical Trial Registry (ChiCTR2300068073)

**Highlights:**

1. It is the first time of our study to find the remarkable elevated effect of intranasal insulin on peripheral and central osteocalcin.
2. Besides, intranasal insulin showed a significantly reduction in the incidence and severity of postoperative delirium in elderly patients undergoing joint replacement, as well as an amelioration in insulin sensitivity.
3. Considering the positive role of osteocalcin played in insulin secretion and brain function maintenance, our findings may suggest that the improvements of peripheral insulin sensitivity and decrease of postoperative delirium are probably associated with elevated peripheral and central osteocalcin levels.
4. Though further studies are needed to confirm, it suggested that osteocalcin is a promising biomarker that involves in the effect of intranasal insulin both in improving peripheral metabolism and central cognition.

## Introduction

Postoperative delirium is a common and life-threatening complication in elderly surgical patients, characterized by acute disorders of attention and cognition (1, 2). As the global average life expectancy increases and the population ages, the incidence of postoperative delirium is increasing year by year (3). The prevention and treatment of delirium remains a difficult problem due to the side effects and limited efficacy of the existing treatment options (4). Recently, intranasal insulin has shown a promising application prospect in the prevention and treatment of perioperative neurocognitive disorders without any side effect (5–7), but its specific mechanism is uncertain.

The protective effect of intranasal insulin on perioperative neurocognitive disorders is inseparable to its improvement of peripheral insulin sensitivity and central cognitive function. At present, it is known that intervention of intranasal insulin could improve the peripheral insulin sensitivity by promoting the glucose uptake and catabolism in tissues and muscles with specific pathway unclear (8). Interestingly, studies have shown that osteocalcin signal in muscles promotes glucose uptake and catabolism, and even in insulin deficiency or insulin resistance state (9, 10). Moreover, osteocalcin levels were related to insulin sensitivity, and elevated osteocalcin levels contributed to the amelioration of insulin sensitivity (11). More importantly, osteocalcin, as an important hormone that tightly linked bone metabolism, energy metabolism and cognitive function together, also plays an important role in the regulation of cognitive function(12). Osteocalcin could cross the blood brain barrier, accumulated at the specific area of brain, and participated in the regulation of cognition(13). The mice lack of osteocalcin showed abnormalities in cognitive function of impaired learning and memory, which were alleviated after osteocalcin supplementation (12). In elderly patients, low osteocalcin was associated with brain microstructural changes and worse cognitive performance(14, 15). However, osteocalcin supplementation can also save age-related cognitive decline in older mice(16). In brief, insulin signal could promote increased levels of osteocalcin (17), and increased osteocalcin appeared to improve insulin sensitivity and cognitive function. Therefore, we hypothesize that intranasal insulin improves central cognitive function and peripheral insulin sensitivity by increasing peripheral and central osteocalcin levels and plays an important role in the prevention of perioperative neurocognitive decline.

The aim of this study was to explore the effect of intranasal insulin on the incidence and severity of postoperative delirium, insulin sensitivity and osteocalcin levels in elderly patients undergoing joint replacement.

## Results

### Patient demographics and baseline characteristics

A total of 318 adults were screened, and 212 elderly patients undergoing selective joint replacement were recruited and randomized into insulin group (n = 106) and placebo group (n = 106). 10 patients in insulin group and 7 patients in placebo group were dropped out for not receiving intervention of intranasal drugs, failing on combined epidural anesthesia or canceled surgery. Finally, 96 in the insulin group and 99 in the placebo group were analyzed in the study, with 145 female and 50 male patients (showed in Figure 1).

**Figure 1.**
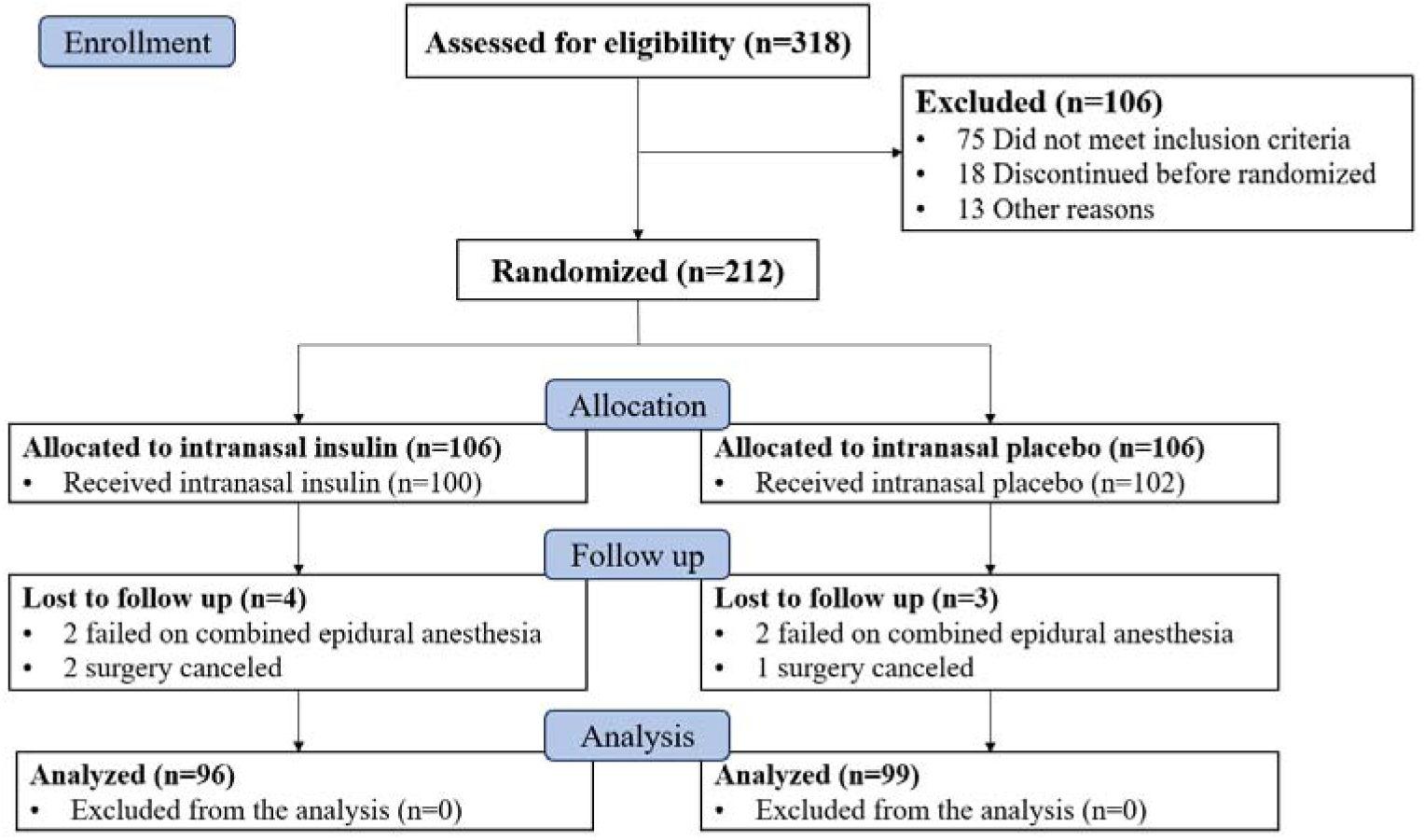
Flowchart of the study design.

Demographic data, comorbidity, surgery data and biomarkers at baseline are shown in Table 1. The demographic data between the two groups were matched including sex, age, BMI, education, MMSE, physical activity, smoking and alcohol habit. The anamnesis of two groups balanced well, including Self-reported cardiac disease, elf-reported or imaging diagnosed stroke/TIA, self-reported hypertension, and Self-reported diabetes. There were no significant differences in surgery surgical type, preoperative fracture, duration of surgery, intraoperative blood loss, visual analogue score between the insulin and placebo group. Before intervention, there were no significant differences in the levels of biomarkers in cerebrospinal fluid and plasma, including ucOC, OC, insulin, glucose and HONA-IR.

**Table 1.** Baseline demographic and clinical characteristics.

| Variable | Overall(n=195) | Insulin ( n=96 ) | Placebo ( n=99 ) |
| --- | --- | --- | --- |
| General data |  |  |  |
| Age (y) | 72.9±6.32 | 72.8±6.0 | 73.0±6.68 |
| Female, n (%) | 145(74.35%) | 72(75%) | 73(76.04%) |
| BMI | 23.46±3.44 | 23.06±3.09 | 23.84±3.72 |
| Education(y) | 7.7±4.58 | 7.4±4.58 | 7.9±4.60 |
| MMSE | 25.38±3.72 | 25.19±3.50 | 25.58±3.93 |
| Physical activity |  |  |  |
| Inactivity | 89(45.6%) | 41(42.7%) | 48(48.5%) |
| 1-2 times/week | 74(37.9%) | 37(38.5%) | 37(37.4%) |
| >2 times/week | 32(16.4%) | 18(18.8.6%) | 14(14.1%) |
| Smoking, n (%) | 20(10.26%) | 12(12.5%) | 8(8.08%) |
| Alcohol, n (%) | 8(4.10%) | 4(4.17%) | 4(4.04%) |
| Self-reported cardiac disease, n (%) | 33(16.92%) | 18(18.75%) | 15(15.15%) |
| Self-reported or imaging diagnosed stroke/TIA, n (%) | 30(15.38%) | 14(14.58%) | 16(16.16%) |
| Self-reported hypertension, n (%) | 98(50.26%) | 56(58.33%) | 42(42.42%) |
| Self-reported diabetes, n (%) | 46(23.59%) | 24(25.26%) | 22(22.22%) |
| Insulin treatment, n (%) | 18(9.23%) | 9(9.47%) | 9(9.10%) |
| Oral hypoglycemic drugs, n (%) | 28(14.36%) | 15(15.79%) | 13(13.13%) |
| Surgery data |  |  |  |
| Surgical type/total hip, n (%) | 103(52.82%) | 56(58.33%) | 47(47.47%) |
| Preoperative fracture, n (%) | 56(28.72%) | 32(33.33%) | 24(24.24%) |
| Duration of surgery (hours) | 4.08±1.30 | 4.01±1.23 | 4.15±1.43 |
| Intraoperative blood loss (ml) | 234.3±211.8 | 238.3±213.5 | 230.6±219.7 |
| Visual Analogue Score | 2.55±1.12 | 2.59±1.28 | 2.47±1.07 |
| Baseline markers |  |  |  |
| UcOC in plasma(ng/mL) | 1.54±0.59 | 1.52±0.58 | 1.56±0.57 |
| TOC in plasma(ng/mL) | 13.36±8.48 | 13.25±8.78 | 13.47±8.92 |
| UcOC/tOC in plasma (%) | 15.94±12.27 | 16.05±10.36 | 15.89±11.53 |
| Insulin in plasma(uU/mL) | 34.09±10.15 | 34.87±10.32 | 33.64±9.52 |
| Glucose in plasma(mmol/L) | 5.26±0.91 | 5.31±0.98 | 5.19±0.76 |
| HOMA-IR | 7.99±2.79 | 8.21±2.95 | 7.75±2.40 |

MMSE, mini-mental state examination, BMI, Body mass index; ucOC, uncarboxylated osteocalcin;
tOC, total osteocalcin; HOMA-IR, Homeostasis model assessment of insulin resistance.

### Effect of intranasal insulin on osteocalcin levels

The differences of osteocalcin levels in plasma and cerebrospinal fluid between two groups at different days after intervention are showed in Table 2. There were significantly differences of ucOC and tOC in cerebrospinal fluid and plasma between insulin and placebo groups on the day of surgery (D_0_), day 1 after surgery (D_1_) and day 3 after surgery (D_3_). After 3 days of intranasal insulin intervention, the ucOC and tOC levels in cerebrospinal fluid were apparently elevated in insulin group at D_0_ showed in Figure 2 (P<0.001, P<0.001). Comparison of osteocalcin levels in plasma at different days after intervention between two groups showed in Figure 3. Plasma tOC levels were also significantly increased after intervention in insulin group at D_0_, D_1_, and D_3_ (P<0.001, P<0.001, P<0.001). Plasma ucOC levels were significantly higher in insulin group than placebo group at D_0_ (P<0.001), and significantly lower in insulin group than placebo group at D_1_, and D_3_ (P<0.001, P<0.001). To compare the difference in the percentage of ucOC to tOC between the two groups and found that the ratio of ucOC to tOC (ucOC/tOC) in insulin group was all significantly lower than that in placebo group on D_0_, D_1_ and D_3_ showed in Table 2 (P<0.001, P<0.001, P<0.001).

**Figure 2.**
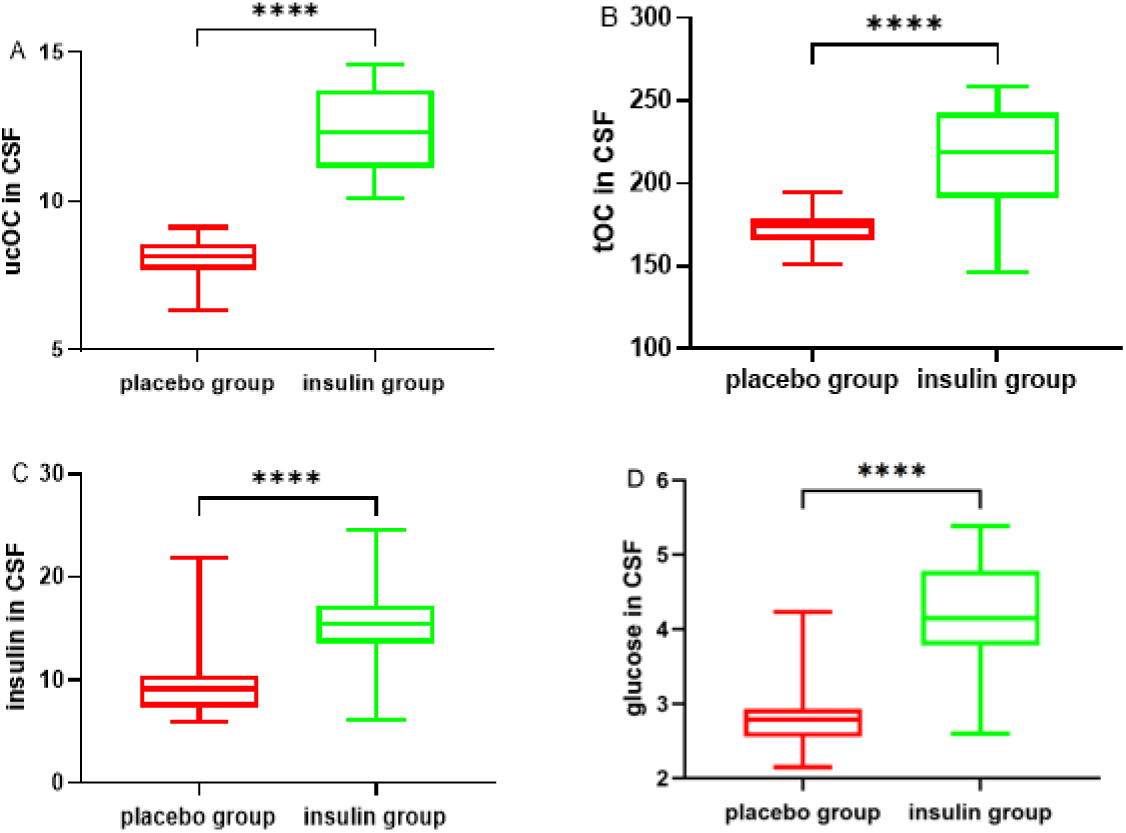
Comparison of biomarkers in cerebrospinal fluid between the two groups. A, the difference of uncarboxylated osteocalcin in cerebrospinal fluid between the two groups; B, the difference of total osteocalcin in cerebrospinal fluid between the two groups; C, the difference of insulin levels in cerebrospinal fluid between the two groups; D, the difference of glucose levels in cerebrospinal fluid between the two groups. **** P<0.001.

**Figure 3.**
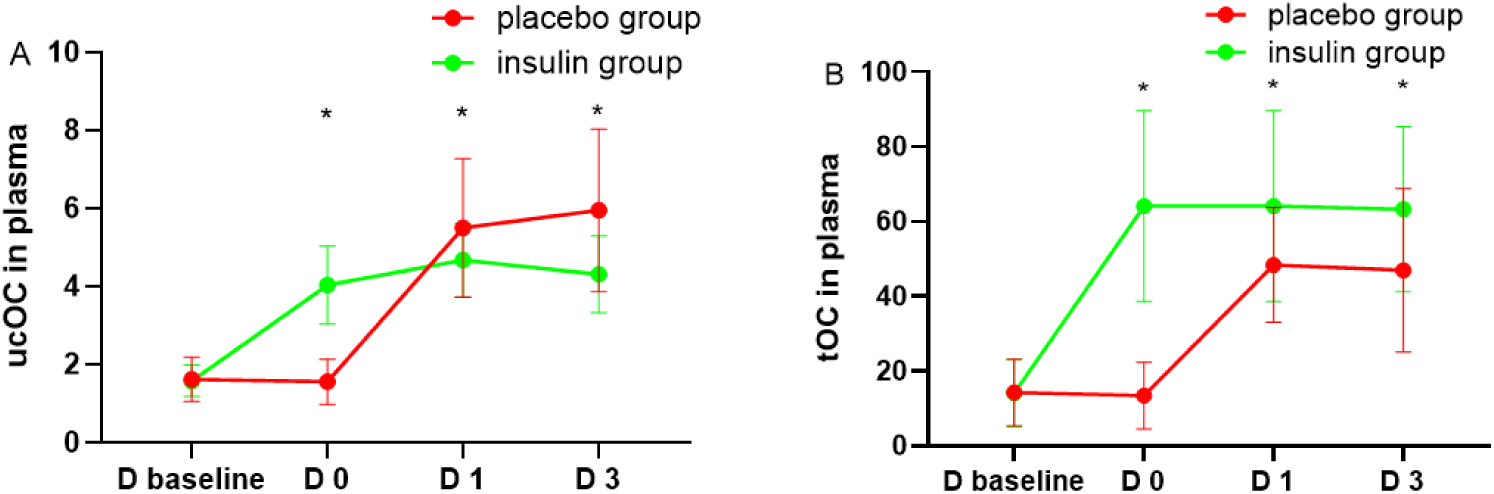
Changes of plasma osteocalcin levels in the two groups. A, the line chart of uncarboxylated osteocalcin concentration in two groups on D_baseline_, D_0_, D_1_ and D_3_. B, line chart of total osteocalcin concentration in two groups on D_baseline_, D_0_, D_1_ and D_3_. D_baseline_, the day before intervention, D_0_, the day of surger, D_1_, the first day after surgery, D_3_, to the third day after surgery. * P<0.05.

**Table 2.** Comparison of the osteocalcin levels between the two groups.

| markers | all ( n=195 ) | placebo<br>group(n=99) | insulin<br>group(n=96) | *P* value |
| --- | --- | --- | --- | --- |
| ucOC in CSF at D0 (ng/mL) | 10.22±2.38 | 8.12±0.61 | 12.38±1.37 | <0.001* |
| ucOC in plasma at D0 (ng/mL) | 2.78±1.48 | 1.57±0.57 | 4.03±1.00 | <0.001* |
| ucOC in plasma at D1 (ng/mL) | 5.10±1.48 | 5.50±1.78 | 4.68±0.94 | <0.001* |
| ucOC in plasma at D3 (ng/mL) | 5.15±1.82 | 5.95±2.08 | 4.31±0.98 | <0.001* |
| tOC in CSF at D0 (ng/mL) | 192.24±31.28 | 171.29±10.60 | 213.85±30.86 | <0.001* |
| tOC in plasma at D0 (ng/mL) | 38.42±31.69 | 13.47±8.92 | 64.14±25.51 | <0.001* |
| tOC in plasma at D1 (ng/mL) | 56.06±17.57 | 48.37±15.37 | 63.99±16.19 | <0.001* |
| tOC in plasma at D3 (ng/mL) | 55.02±23.39 | 46.98±21.84 | 63.31±22.11 | <0.001* |
| ucOC/tOC in plasma at D0(%) | 11.54±9.97 | 16.11±12.27 | 6.82±1.75 | <0.001* |
| ucOC/tOC in plasma at D1(%) | 9.86±3.88 | 11.97±3.95 | 7.69±2.30 | <0.001* |
| ucOC/tOC in plasma at D3(%) | 10.87±5.94 | 14.32±6.45 | 7.31±2.02 | <0.001* |

ucOC: uncarboxylated osteocalcin; tOC: total osteocalcin; CSF: cerebrospinal fluid; D0: the day of
surgery; D1: the first day after surgery; D3: The third day after surgery.

### Effect of intranasal insulin on insulin, glucose and HOMA-IR

Peripheral HOMA-IR in insulin group was nearly the same to that in placebo group at D_0_ (P=0.669), and it was markedly decreased in insulin group at D_3_ (P=0.017) showed in Figure 4.

**Figure 4.**
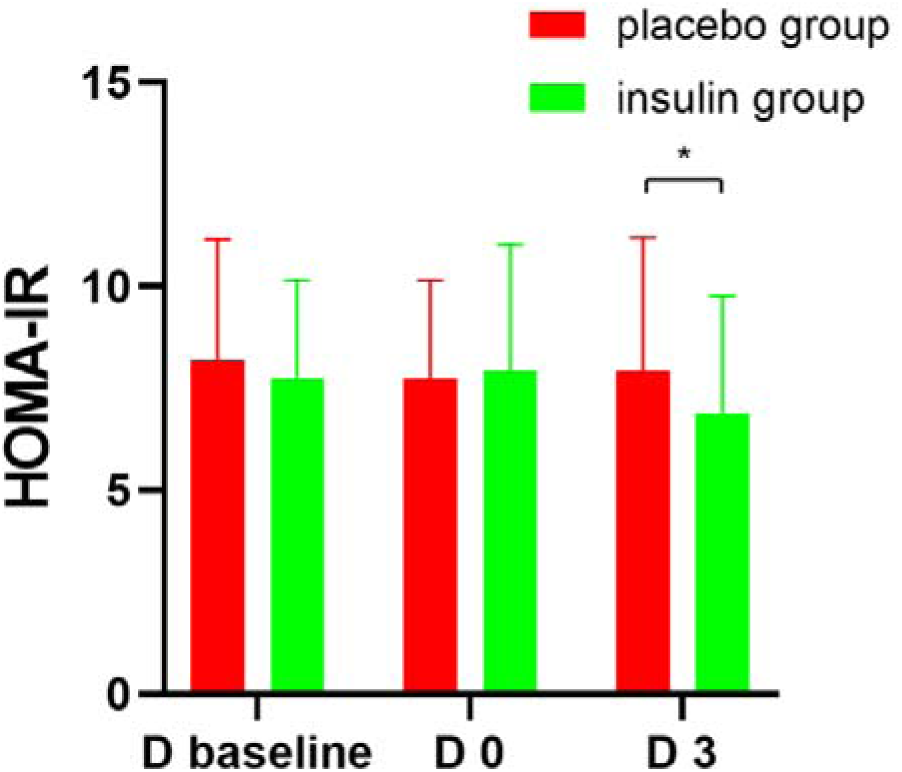
Comparison of HOMA-IR between the two groups. It shows the comparison of HOMA-IR between the two groups at D_baseline_, D_0_, and D_3_. *P<0.05.

In plasma, there were no obvious difference in the glucose levels between two groups after intervention at D_0_, D_1_, and D_3_. And the levels of insulin almost stayed the same after intervention at D_0_, but it in insulin group was visibly decreased compared to placebo group at D_3_. In cerebrospinal fluid, the insulin and glucose levels were apparently elevated in insulin group after 3 days of intranasal insulin intervention at D_0_ showed in Figure 2 (P<0.001, P<0.001).

### Effects of intranasal insulin on the incidence and severity of POD

A total of 195 patients finished the trial, all of whom completed the delirium assessment. Among the participants, a total of 31 patients (15.90%) developed postoperative delirium, among which the incidence of POD in the insulin group (8/96, 8.33%) was significantly lower than that in the placebo group (23/99, 23.3%) showed in Table 3 (P=0.004). The incidence of POD was 29.0% (9/31) for hypoactive type, 51.6% (16/31) for hyperactive type, and 19.4% (6/31) for mixed type. The severity of delirium was mainly mild to moderate, and the median duration was 1.0 days in the insulin group and 2.0 days in the placebo group, respectively. The mean peak-DRS in insulin group was significantly lower than that in placebo group showed in Table 3 (P<0.001).

**Table 3.** Comparison of postoperative delirium incidence and severity between two groups.

| Variable | Overall | Insulin group | Placebo group | *P* value |
| --- | --- | --- | --- | --- |
| POD | 31(15.90%) | 8(8.33%) | 23(23.23%) | 0.004* |
| Peak-DRS | 8.01±6.0 | 6.01±3.5 | 9.94±7.1 | <0.001 |

POD, postoperative delirium; Peak-DRS, peak values of the Delirium Rating Scale-98.

## Discussion

In this randomized, double-blind, placebo-controlled clinical trial, intranasal insulin remarkably elevated osteocalcin levels in cerebrospinal fluid and plasma, improved peripheral insulin sensitivity, and significantly reduced the incidence and severity of postoperative delirium in elderly patients undergoing joint replacement. This not only found that intranasal insulin can significantly increase peripheral and central osteocalcin levels, but also suggested that osteocalcin may be closely related to the mechanism of intranasal insulin in improving central cognitive function and peripheral insulin sensitivity.

The significantly improvement of intranasal insulin on short-term memory and cognitive function have been widely validated and recognized (18), although in a long-term intervention study of it showed without obvious cognitive or functional benefits in patients with mild cognitive impairment or Alzheimer’s disease (19). Encouragingly, in a large number of animal studies and some clinical studies, intranasal insulin has shown a promising application prospect in the transient and reversible perioperative neurocognitive impairment (5–7, 20, 21). Our results supported the prominent effect of intranasal insulin in the prevention and treatment of postoperative delirium and explored its possible mechanism.

The mechanism of intranasal insulin in improving memory and cognition is complex and diversified. It was showed in our findings that the mean central glucose concentration in insulin group (4.57mmol/L) was significantly higher than that in placebo group (2.83mmol/L) after intranasal insulin. According to the verification from imaging, low glucose metabolism in the specific region of brain has been reported to be the main manifestation of consciousness and cognition impairment (22). Intranasal insulin could increase the central brain energy levels in study on healthy subjects (23). It could elevate glucose levels in several regions of brain under exam of positron emission tomography, thus linking improvements in cognitive function through intranasal insulin to restoration of glucose metabolism in these specific brain regions (24). Hence, the mechanism of intranasal insulin might be associated with the improvement of central energy substance level and glucose metabolism. Furthermore, intranasal insulin notably risen central insulin levels in our results, which could be related to the amelioration of brain insulin resistance. Brain insulin resistance, as the pathological feature of metabolic and cognitive disease, is characterized by decreased brain insulin uptake and sensitivity (25). The underlying mechanisms might include insulin resistance at the blood-brain barrier, resulting in deficiency of central insulin (26). Therefore, intranasal insulin could ameliorate cognitive function through directly elevating central insulin levels and improving brain insulin resistance. Overall, these finding were consistent with the known mechanism of intranasal insulin.

Of note, intranasal insulin rapidly increased central and plasma osteocalcin levels in our findings. Osteocalcin, as a bone-derived hormone, is necessary to promote normal brain development and function, and recently has been described as sufficient to reverse the cognitive manifestations of aging (27). As early as embryonic development, maternal osteocalcin affects fetal brain development for its favor to promote fetal neurogenesis and prevent neuronal apoptosis (13). Osteocalcin gene knockout mice showed significantly increased anxiety-like behavior and severe deficits in learning and memory, which could be fully corrected by central supplementation of osteocalcin (12). The increase of serum osteocalcin correlated with improvement in cognitive dysfunction after the therapy of liraglutide in diabetic rats, supporting the protective effect of elevated peripheral osteocalcin on cognition (28). After middle age, circulating osteocalcin declined, contributing in part to age-related cognitive decline. Radiologically, lower serum osteocalcin levels were associated with brain microstructural changes and poorer cognitive performance in elderly patients (14). In old mice, central osteocalcin supplementation is a necessary and sufficient condition to correct age-related cognitive decline, as strong evidence of animal models that elevated central osteocalcin improves cognition (16). Despite lack of clinical evidence that elevated central osteocalcin improves cognition, the incidence of postoperative delirium was significantly reduced after the remarkable elevation of central and peripheral osteocalcin under intervention of intranasal insulin in our findings. This might be indirect evidence that central and peripheral osteocalcin elevations improve cognition, and also suggested that osteocalcin was probably involved in the mechanism of nasal insulin to improve cognition.

There has been increasing evidence that intranasal insulin can rescue glucose homeostasis and insulin sensitivity (29), with the underlying molecular mechanism uncertain. It is showed that intranasal insulin administration can rapidly enhance peripheral insulin sensitivity through increasing suppression of endogenous glucose production and stimulating peripheral glucose uptake (30), while peripheral insulin levels stayed almost constant. It suggested that there may be other molecule besides plasma insulin directly promoting peripheral glucose uptake. Recently, osteocalcin has been proved with the ability of promoting glucose uptake of muscle tissue in both tissue and animal models (9, 31). And that, there is a bidirectional positive feedback mechanism between osteocalcin and insulin (17). Insulin signal in osteoblast could promote the secretion of osteocalcin, and in return, osteocalcin could promote the secretion of insulin and improve the insulin sensitivity. Hence, improving insulin signaling in osteoblasts might help increase circulating osteocalcin levels and further improves insulin sensitivity by promoting insulin secretion and promoting glucose uptake by tissues. As existing evidence to support this view, it showed that plasma osteocalcin was positive related with insulin sensitivity (32, 33) and raised plasma osteocalcin levels was linked to the improvement of insulin sensitivity (11). It is surprisingly that intranasal insulin promptly increased the plasma total osteocalcin level and subsequently improved peripheral HOMA-IR in our results. Therefore, osteocalcin might be related to the molecular mechanism of intranasal insulin on improving peripheral HOMA-IR and insulin sensitivity. Though more researches need to confirm the potential role of osteocalcin played on the mechanism of intranasal insulin, osteocalcin seems to be a promising molecular that could explain the effect of intranasal insulin both in peripheral metabolism and central cognition.

The effect of uncarboxylated osteocalcin in plasma on cognitive function was contradictory. Serum ucOC level seemed to be a protective effect to cognition and was positively related with cognitive impairment in rats or males with type 2 diabetes (34, 35). While, elevated levels of plasma uncarboxylated osteocalcin were associated with impaired cognitive function in community-dwelling older adults (36). In our previous study, preoperative uncarboxylated osteocalcin in cerebrospinal fluid were an independently risk factors to postoperative delirium in elderly patients underwent joint replacement (37). In the results of this study, the level of uncarboxylated osteocalcin in insulin group was significantly lower than that in placebo group after surgery, although intranasal insulin elevated the uncarboxylated osteocalcin levels before surgery. Beyond that, intranasal insulin distinctly reduced the ratio of uncarboxylated osteocalcin to total osteocalcin. Osteocalcin is converted from uncarboxylated osteocalcin into carboxylated osteocalcin by key rate-limiting enzymes (38), so low level of uncarboxylated osteocalcin, especially decreased ratio of uncarboxylated osteocalcin to total osteocalcin, probably indicated a better status of vitamin K. Vitamin K, as a vital cofactor, has shown a protective effect on insulin sensitivity and cognitive function (39). Therefore, the reduced uncarboxylated osteocalcin and ratio of uncarboxylated osteocalcin to total osteocalcin in insulin group were probably reflected the amelioration of insulin sensitivity and cognition by intranasal insulin.

In conclusion, our study found that intranasal insulin significantly reduced the incidence and severity of postoperative delirium, and improved central and peripheral osteocalcin levels and insulin sensitivity in elderly patients with joint replacement. This study not only expanded the application of intranasal insulin in the prevention of perioperative neurocognitive dysfunction, but also suggested the potential role of osteocalcin played underlying the mechanism of intranasal insulin in rescuing peripheral metabolism and central cognition.

## Materials and Methods

### Participants and Study Design

Recruitment took place between 1 December 2022 and 18 July 2023 from the Third Xiangya Hospital of Central South University and was stopped for futility in accordance with the DSMB. Eligible patients were those aged 65 years and older, American Society of Anesthesiologists (ASA) physical status I to III, scheduled for elective joint replacement surgery under combined epidural anesthesia and stayed in the hospital after surgery for more than 5 days. Additional inclusion criteria were that the participants must be able to communicate with the researcher and complete the relevant scale assessment (no serious visual impairment or hearing problems). The exclusion criteria were any patients with preexisting psychiatric or neurological disease (e.g., intracranial tumors), surgery within the past year, and a baseline Mini-Mental State Examination (MMSE) score less than 23 (40), or other conditions that the attending physician or investigator deems inappropriate to participate in the study (the patient has a serious nasal disease that affects the efficacy of intranasal administration). To fully understand the geriatric characteristics of the participants, a comprehensive geriatric assessment was performed on all patients in Appendix 1.

### Randomization and blinding

After giving consent, eligible patients were randomized using a random number (in a 1:1 ratio) generated on a computer program (SAS 9.2 software) and assigned to receive nose sprays in the morning and night with either dosage of 40 IU insulin detemir (1 ml) or placebo (1 ml normal saline) twice a day from the 3rd day before surgery to the 5th day after surgery for 8 days (41). Insulin detemir (Levemir®; Novo Nordisk, Novo Alle, Denmark) was repackaged in a set of nasal spray bottles (Aeropump, Hochheim am Main, Germany) in our Department of Clinical Pharmacy Experimental Center. Equal volumes of normal saline were provided in the same packaging bottle for the placebo group. Masked drug was provided by a pharmacist, who was independent of investigators and clinicians. The results of randomization were sequentially sealed in numbered envelopes until the end of the study. In emergency of clinical work (such as an accident or rapid deterioration of the patient’s condition), the professor involved in clinical treatment can immediately demand to see the status of the treatment group in order to adjust or interrupt the administration of the study drug to the subject if necessary. All of these incidents are recorded in the subject’s report file.

### Anesthesia Procedures

During the entire perioperative period, all clinical management followed recognized clinical practice. All patients were given combined epidural anesthesia. Lumbar anesthesia drugs are 1.5ml 1% ropivacaine +0.025mg fentanyl +0.5ml 10% glucose +0.5ml cerebrospinal fluid. Epidural drug supplementation 2 to 3h after the administration of lumbar anesthesia, 4ml 2% lidocaine was first given as an experimental dose according to the anesthesia level, and then 5ml 0.75% ropivacaine injection was added every 1.5 to 2h according to the anesthesia level. All subjects did not receive preoperative medication (such as atropine or diazepam). Other aspects of management (blood pressure indicators, use of vasoactive drugs) are at the discretion of the attending anesthesiologist. Avoid medications known to affect cognition (such as benzodiazepines or dexmetropine). All subjects were treated with patient control analgesia (PCA). 150ug sufentanil + 8mg ondansetron were diluted to 150ml with normal saline at a speed based on height, weight, pain threshold, etc. The VAS score of all patients was controlled to ≤3, and the treatment regimen was the same (all patients were given intravenous dezocine injection). All the conditions of all patients are recorded in the case report.

## Outcome measures

### Measurements of osteocalcin, insulin, glucose, and insulin sensitivity

Peripheral blood was acquired from 6 to–8La.m. on the day before intervention (D_baseline_), the day before surgery (D_0_), the first day after surgery (D_1_), and the third day after surgery (D_3_), and obtained plasma was immediately centrifuged at 3000 rpm for 10 min at 4°C. CSF was acquired in the immediate preLoperative period during induction of spinal anesthesia (PREOP). All the samples were immediately frozen at −80°C. EnzymeLlinked immunosorbent assays were used to measure the biomarkers. The concentrations of total osteocalcin (tOC), uncarboxylated osteocalcin (ucOC) and insulin in plasma and in CSF were all measured using a solid□phase enzyme-linked immunosorbent assay (ELISA) kit (Jiancheng, Nanjing, China). The concentrations of glucose in plasma and in CSF were measured using a glucose assay kit (Jiancheng, Nanjing, China) through the glucose oxidase method. Detection limits of biomarkers were different, and all detected results were included within the limits. Regarding the repeatability of the ELISA kit, the coefficient of variation was less than 10% both within and between the plates.

Insulin sensitivity was estimated by the homeostasis model assessment of insulin resistance (HOMA-IR) (42). The HOMALIR was calculated using the following formula, Fasting Insulin × Fasting Glucose/22.5. A baseline HOMALIR index greater than 2.6 was predefined arbitrarily as insulin resistance. According to our previous study (6), although intranasal insulin improves insulin sensitivity quickly (within a few hours), the time of significant changes in HOMA-IR is relatively late. Therefore, the detection time of HOMA-IR is selected as the 4th and 7th days after intervention (namely at D_0_ and D_3_).

### Measurements of the incidence and severity of postoperative delirium

The secondary outcome was the incidence and severity of postoperative delirium, determined by CAM and DRS-98 (43). For 5 days after surgery, the patient was assessed by two trained visitors once a day in the morning and evening for the CAM scale and the Visual Pain Scale, and a Delirium Rating Scale-98 (DRS) was performed to assess the severity of delirium. The patient’s delirium was assessed by the experimental delirium evaluator between 8:00-10:00 AM and 19:00-21:00 PM, and evaluated twice a day. Possible episodes of delirium outside the assessment time were evaluated by interviewing the patient’s companion and ward nurse, as well as nursing records. The clinical research assistants who performed delirium assessments in this study were well trained and underwent quality control procedures. POD was evaluated using The Confusion Assessment Method (CAM). Four clinical criteria were used to diagnose POD, (1) acute onset and fluctuating course of disease; (2) inattention; (3) Confused thinking; (4) The level of consciousness changes. To define delirium (1) and (2) must both be satisfied, plus (3) and/or (4). The severity of the POD is determined using a score on the Delirium Rating Scale-98 (DRS), with higher scores indicating more severe delirium. All patients were divided into postoperative POD group (POD) and postoperative delirium-free group (non-POD) according to whether they developed delirium or not.

In our study, we conducted delirium assessments twice a day in the morning and twice a day during working hours, which also means that delirium occurring at night may have gone undetected. However, we followed up with family members and nurses to find out about the patient’s mental state during this period. The shedding rate of the primary outcome was very low, so we consider the results of this study to be credible in assessing postoperative delirium.

### Ethics

Ethical approval for this study (ethical code, K22051) was provided by the Ethical Committee of the Third Xiangya Hospital of Central South University (Chairperson Prof Zhiying Su) on 20 October 2022(address, Ethics Committee, 138 Tongzipo Road, Yuelu District, Changsha City, China).

### Standard protocol approvals, registrations, and patient consent

We conducted this randomized, double-blind, placebo-controlled clinical trial at the Third Xiangya Hospital of Central South University between December 2022 and July 2023. The study was registered with the Chinese Clinical Trial Registry (ChiCTR2300068073). The study was conducted following the Consolidated Standards of Reporting Trials (CONSORT) reporting guideline 2010. Written consent was obtained from patients or a legal representative. The study was conducted under local institutional review board supervision.

## Statistics

Sample Size. Our previous prospective cohort study showed that the incidence of POD 5 days in elderly patients after joint replacement surgery was 23.3% (37), the sample size calculation used a two-sided design at a significance level of 5% (α = 0.05) and a power of 95% (1-β = 0.95) assuming an incidence of 23% and indicated that 106 patients per group would be needed to detect a 60% decrease in the incidence of POD. We finally recruited 106 patients in each group. After a formal review requested by the data monitoring and ethics committee, the sample size was revised to reflect a lower attrition rate than anticipated.

For the primary outcome, the differences in osteocalcin levels and insulin sensitivity between the two groups were analyzed by T-test or rank sum test according to whether the data conformed to normal distribution. For secondary outcomes, the difference in delirium incidence between the two groups was analyzed by Chi-square test. To compare the difference of delirium severity, the difference of peak DRS value between the two groups was analyzed by rank sum test.

SPSS (SPSS Inc., Chicago, IL, USA) 22.0 was used for data analysis, where values of P < 0.05 were considered statistically significant. Graphics were created with GraphPad Prism (GraphPad software, San Diego, CA, USA) 8.0 for Windows.

## Data Availability

All data produced in the present study are available upon reasonable request to the authors

## Acknowledgements

We thank the parents and patients for making this study possible. Thanks to the Department of Anesthesiology, Orthopectics and the Central laboratory for their help.

## Author contributions

YM conceived and helped design the study. YM, LG, HX, LX and XC acquired the data. YM, OYW, ZL and HX analyzed and interpreted the data. All the authors made substantial contributions to drafting or revising the article critically for important intellectual content and approved the final version to be published. Each author takes public responsibility for it.

