## Appendix 1 for "Effect of intranasal insulin on osteocalcin levels and postoperative delirium in elderly patients undergoing joint replacement"

Appendix 5. Comprehensive geriatric assessment.

| Geriatric condition | All（195） | Insulin（96） | Placebo（99） |
| --- | --- | --- | --- |
| **Comorbidity** |  | | |
| 1-2 types | 136(69.7%) | 63(65.6%) | 73(73.7%) |
| 3+ types | 59(30.3%) | 33(34.4%) | 26(26.3%) |
| **Medicine** |  | | |
| 0-4 types | 153(78.5%) | 71(74.0%) | 82(82.8%) |
| 5+types | 42(21.5%) | 25(26.0%) | 17(17.2%) |
| **BMI** |  | | |
| Undernutrition (BMI<20 kg/m²) | 22(11.3%) | 14(14.6%) | 8(8.1%) |
| Normal (20≤BMI≤30 kg/m²) | 156(80.0%) | 73(76.0%) | 83(83.8%) |
| Overweight (BMI >30 kg/m²) | 17(8.7%) | 9(9.4%) | 8(8.1%) |
| **Physical activity** |  | | |
| Inactivity | 89(45.6%) | 41(42.7%) | 48(48.5%) |
| 1-2 times/week | 74(37.9%) | 37(38.5%) | 37(37.4%) |
| ≧3 times/week | 32(16.4%) | 18(18.8.6%) | 14(14.1%) |
| **MMSE** | 25.38±3.72 | 25.19±3.50 | 25.58±3.93 |

Comorbidity, including hypertension, diabetes, cardiac disease, cerebral infarction and chronic obstructive pulmonary disease; BMI, Body Mass Index. MMSE, Mini-mental State Examination.
